# Diabetes mellitus among older people in rural Sidama, Ethiopia: a two-step community-based cross-sectional study

**DOI:** 10.1101/2025.08.19.25333977

**Authors:** Desalegn Tsegaw Hibstu, Melaku Haile Likka, Hiwot Abera Areru, Betelhem Eshetu Birhanu, Bernt Lindtjørn

## Abstract

**Objective:** To determine the prevalence of undiagnosed diabetes and identify associated factors using a two-step diagnostic method combining fasting blood sugar with confirmatory glycated haemoglobin A1c (HbA1c) testing among older adults in rural Sidama, Ethiopia.

**Design:** A community-based cross-sectional design was conducted from April 1 to July 31, 2024. Data were collected through a census of adults aged ≥ 45 years using a pretested WHO-STEPwise questionnaire. Physical and biochemical tests were performed following standard protocols. Data were analysed using Stata version 17.

**Setting:** Selected rural kebeles of Shebedino district, Sidama, Ethiopia.

**Participants:** 2,875 adults aged ≥ 45 years.

**Primary outcome measures:** Undiagnosed diabetes confirmed by haemoglobin A1c levels (≥48 mmol/mol).

**Results:** The prevalence of undiagnosed diabetes confirmed by haemoglobin A1c was 1.2% (35 of 2,871; 95% CI: 0.9 – 1.7%). Previously diagnosed diabetes was found in in 0.5% (14 of 2,875; 95% CI: 0.3% - 0.8%). The total diabetes prevalence, confirmed by haemoglobin A1c or prior diagnosis, was 1.7% (49 of 2871; 95% CI: 1.3% - 2.3%). Nearly half, 46.2% (1,327 of 2,875), were undernourished. Advanced age (β = 0.18; 95% CI: 0.06, 0.30, p = 0.004), estimated annual income (β = 0.14; 95% CI: 0.01, 0.27; p = 0.039), and waist-to-body mass index (β = 0.08; 95% CI: 0.01, 0.16; p = 0.032) were significantly associated with elevated fasting blood sugar levels.

**Conclusion:** The prevalence of undiagnosed diabetes in this rural setting was low. Increasing age, higher income, and waist-to-body mass index were associated with elevated fasting blood sugar. Routine community-based diabetes screening, health education, and nutrition-focused interventions are recommended to sustain the low burden and address undernutrition.

**Strengths and limitations of this study:**

- A two-step diagnostic approach (fasting blood sugar and confirmatory HbA1c) enhanced the diagnostic accuracy compared with fasting blood sugar alone.
- Census-based sampling of a large rural population aged 45 years and above improved representativeness and reduced selection bias.
- Standardised tools (WHO-STEPwise protocol) and trained field teams were used to ensure data quality.
- Diabetes types were not differentiated with additional biomarkers (e.g., autoantibody or C-peptide testing), potentially causing misclassifications.
- Self-reported behaviours (smoking, khat chewing, and alcohol use) may be prone to recall or social desirability bias.

## Introduction

Diabetes mellitus is a chronic metabolic disorder characterised by persistent hyperglycaemia resulting from defective insulin secretion by pancreatic β-cells and reduced responsiveness of insulin-sensitive tissues [1]. There are two main types of diabetes: type 1 and type 2. Type 2 diabetes mellitus, which accounts for over 90% of all diabetes mellitus cases globally, has multifactorial causes, with behavioural, genetic, and environmental dimensions [2–4].

Common risk factors such as obesity, physical inactivity, ageing, family history, poor diet, smoking, and alcohol consumption contribute to the disease’s burden [3, 5]. In contrast, individuals experiencing chronic malnutrition frequently exhibit a lower prevalence of diabetes, which is likely attributed to reduced calorie intake and decreased fat accumulation [6–8]. While diabetes has traditionally been associated with urban and high-income settings, recent evidence has shown it is also increasing in rural and low-resource regions, including Sub-Saharan Africa [4].

Globally, type 2 diabetes affects more than 500 million adults (aged 20-79 years), with this figure projected to rise to 643 million by 2030 and 783 million by 2045 [4]. Ethiopia is suffering from the triple burden of communicable diseases, non-communicable diseases, and injuries [9]. Non-communicable diseases account for 37.5% of the disease burden and cause two in five deaths, with diabetes contributing about 3% of all cases [10–12]. Nationally, the prevalence of diabetes mellitus varies widely, from 2% in Tigray to 14% in Dire Dawa [13]. A recent community-based study in the town of Hawassa, Sidama, Ethiopia, showed a prevalence of 14.4% [14]. Additionally, a significant proportion of cases in Ethiopia remain undiagnosed, ranging from 5.8% to 10.2% [15, 16].

Despite the growing recognition of diabetes as a public concern, particularly in rural areas, most studies in Ethiopia have relied primarily on fasting blood sugar for diagnosis [17]. While fasting blood sugar provides only a short-term snapshot of glycaemic control, glycated haemoglobin offers a more reliable measure of long-term blood glucose levels over two to three months [18]. However, few Ethiopian studies have incorporated HbA1c as a confirmatory diagnostic tool, with most conducted in urban settings, such as Addis Ababa, among patients attending diabetic clinic follow-ups [19].

Many diabetic patients in Ethiopia have never undergone HbA1c testing, despite international recommendations supporting its use. This diagnostic gap shows an important limitation in previous diabetes research and control efforts, particularly in underserved rural areas. Due to the lack of HbA1c testing in rural Ethiopia, particularly in community-based studies, a significant knowledge gap remains regarding the true burden of diabetes in Ethiopia [1].

Therefore, the objective of this study was to measure the prevalence of undiagnosed diabetes mellitus in rural Sidama, Ethiopia, using a community-based cross-sectional study design, with HbA1c testing employed as a confirmatory method.

## Methods

### Study design

A community-based cross-sectional study was conducted from April 1 to July 31, 2024, to assess the prevalence of undiagnosed diabetes among older adults in rural Sidama, Ethiopia.

Diabetes mellitus primarily includes type 1, caused by autoimmune β-cell destruction and resulting in absolute insulin deficiency, and type 2, characterised by progressive β-cell insulin dysfunction and insulin resistance [1, 2]. This study focuses on type 2 diabetes, which is more prevalent among older adults, particularly in low-resource settings. Type 1 and other forms were not investigated using different tests, such as autoantibody testing and C-peptide analysis due to their lower expected prevalence in this population [1, 4, 20].

A two-step diagnostic approach was used: initial screening with fasting blood sugar levels (≥ 7 mmol/L), followed by confirmation with haemoglobin A1c testing (≥ 48 mmol/mol). This study is part of a broader initiative to improve non-communicable disease detection and management among older adults in rural Sidama National Regional State [21].

### Study setting

The study was conducted in the Sidama region, Ethiopia. The Sidama region comprises 536 rural kebeles *(the lowest administrative unit, averaging 5,000 residents*), 30 rural districts, and six town administrations, with an estimated population of 4.3 million as of 2020. The region is served by 18 hospitals, 137 health centres, and 553 health posts [22, 23].

Shebedino district, selected purposely for this study, is one of the 36 districts in the Sidama region and has an estimated population of 204,618. It comprises 23 kebeles, one general hospital, six primary healthcare units (*a primary healthcare unit comprises a network of community health posts, health centres, and primary hospitals, forming the backbone of Ethiopia’s healthcare system)*, and 23 health posts. The study was conducted in four kebeles: Gonowa Gabalo, Dobetoga, Howolso, and Gobe Hebisha, located within the catchment area of the Dobe Toga Primary Healthcare Unit, one of the six primary healthcare units in the district. These kebeles collectively house 38,874 residents across 7,933 households. The majority of the population relies on subsistence farming, with *Ensete ventricosum* serving as the primary dietary staple. The Dobe Toga Primary Healthcare Unit delivers maternal and child health services, outpatient care, laboratory, pharmacy, and emergency care services to its catchment population [24].

### Study Participants

The study included all adults aged ≥45 years residing in selected kebeles of Shebedino district, Sidama region, Ethiopia. Individuals were considered eligible if they had resided in the area for at least six months prior to data collection. A complete census was undertaken to identify eligible participants.

### Study variables

The primary outcome was undiagnosed diabetes, identified using haemoglobin A1c testing. The secondary outcome was previously diagnosed diabetes, defined by ≥2 of the following: (1) self-reported diagnosis by a healthcare professional, (2) elevated fasting blood sugar, and (3) current use of anti-diabetic medications, confirmed by visual inspection.

Diabetes was defined per American Diabetes Association criteria: (1) glycated haemoglobin A1c ≥ 48 mmol/mol, performed in a laboratory using National Glycohaemoglobin Standardisation Program (NGSP) certified method, standardised to the Diabetes Control and Complications Trial (DCCT) assay; (2) fasting blood glucose ≥7 mmol/L; (3) 2-hour plasma glucose ≥ 11.l mmol/L after a 75 gram oral glucose load; or (4) random blood sugar ≥ 11.1 mmol/L in individuals with classic hyperglycaemia symptoms [1].

Fasting blood sugar and oral glucose tolerance tests require strict pre-test conditions due to short-term glucose fluctuations. Fasting blood sugar is often measured via capillary blood with a glucometer, while the oral glucose tolerance test involves two timed venous blood draws: before and two hours after the glucose load. Random blood sugar can be taken at any time of the day and is useful for symptomatic individuals. However, all three glucose-based tests are susceptible to variations due to factors such as food intake, stress, illness, and sample handling. Fasting blood sugar reliability depends on a validated glucometer [1, 25].

Therefore, this study used HbA1c as the primary diagnostic tool. HbA1c reflects average blood glucose over 2-3 months and is unaffected by recent meals, activity, or acute illness. It requires no fasting or timed sampling and offers better analytical stability and lower biological variability. These advantages, together with its standardised laboratory methods, made HbA1c a practical and reliable choice for diagnosing diabetes in this rural population of southern Ethiopia [1, 26].

The exposure variables included socio-demographic, economic, behavioural, and clinical factors. The demographic and economic variables covered sex (female and male), age (≥45 years), marital status (single and married), educational level (no formal education, and attended primary education or above), occupational status (farmer, housewife, and other), and travel time to health facility (≤30 minutes and >30 minutes). The wealth index was constructed via principal component analysis from household assets (e.g. type of housing materials, land ownership, crop yield, bank account, and solar light access) and categorised into five levels: poorest, poorer, middle, richer, and richest [27, 28].

Behavioural factors included tobacco use, khat chewing, alcohol intake, fruit and vegetable consumption, and physical activity (vigorous and moderate, each recorded as yes or no).

Anthropometric measurements included height, weight, waist circumference, body mass index (BMI), waist-to-height ratio, and waist-to-body mass index ratio. All the measurements were taken using standard procedures. Height was measured with a stadiometer, with the participants standing barefoot, upright, with their heels, buttocks, and head touching the vertical surface, to the nearest 0.1 centimetre. Weight was measured using a calibrated digital scale placed on a flat surface, with participants wearing light clothing and no shoes, standing still with weight evenly distributed, to the nearest 0.1 kilogram. Body mass index was computed by dividing weight in kilograms by height in metres squared (kg/m^2^). It was categorised as underweight (<18.5), normal weight (18.5–24.9), overweight (25–29.9), or obese (≥30). Waist circumference was measured at the midpoint between the lower margin of the last palpable rib and the top of the iliac crest, using a non-stretchable tape held horizontally and snugly around the waist at the end of normal expiration. Hip circumference was taken at the widest part of the buttocks, also using a non-stretchable tape held horizontally. To assess central obesity relative to overall body size, waist-to-body mass index was computed by dividing waist circumference in centimetres by the body mass index (kg/m²). The waist-to-height ratio was calculated by dividing waist by height, both in centimetres [2, 29].

### Outcome measurement and laboratory analysis

In this study, the confirmation of the outcome variable followed a two-step diagnostic criterion: fasting blood sugar screening and haemoglobin A1c confirmation. Participants were contacted a day before data collection and instructed to fast overnight for at least eight hours. The following morning, before 8:00 a.m., capillary blood samples were collected via finger prick using a Sinocare ISO9001 certified glucometer equipped with automatic calibration that self-adjusts to each test strip. Participants with elevated fasting blood sugar values ≥ 7 mmol/L were referred the next day to Dobe Toga Primary Healthcare Unit for HbA1c confirmatory testing.

To measure the HbA1c, 5 ml of venous whole blood was collected into ethylene diamine tetraacetic acid tubes, which served as anticoagulant to preserve red blood cells. A Trained medical laboratory technician performed the blood collection using standard aseptic procedures. After collection, the tubes were gently inverted 8-10 times to mix blood and anticoagulant, and each tube was immediately labelled with the participant’s unique identifier.

After sample collection at the rural primary healthcare unit, the blood samples were transported in a cold box to Hawassa University Comprehensive Specialised Hospital Laboratory. Upon arrival, the samples were inspected for proper labelling and condition before being stored in a monitored refrigerator, following the guidelines of the Cobas 6000 clinical chemistry analyser (Roche Diagnostics, Mannheim, Germany) for HbA1c sample stability [30]. Testing was conducted using the Cobas 6000, which employs the turbidimetric inhibition immunoassay (TINIA) method to quantify the proportion of glycated to total haemoglobin and expresses the result as mmol/mol. The Cobas 6000 system is standardised to both the International Federation of Clinical Chemistry and Laboratory Medicine (IFCC) and the National Glycohaemoglobin Standardisation Program (NGSP) guidelines [31].

### Sample size and sampling procedure

The initial sample size was estimated using OpenEpi version 3, assuming a 50% prevalence of undiagnosed diabetes, a 95% confidence interval, a 3% margin of error, and a design effect of 2. A 10% non-response was added to account for, resulting in a sample size of 2,371 participants. However, since a census was conducted, the final number of participants included in the study was 2,875 adults aged ≥45 years. For this study, Shebedino district was purposely selected, and no sampling technique was applied due to the census approach. The study flow is indicated in Figure 1.

**Figure 1.**
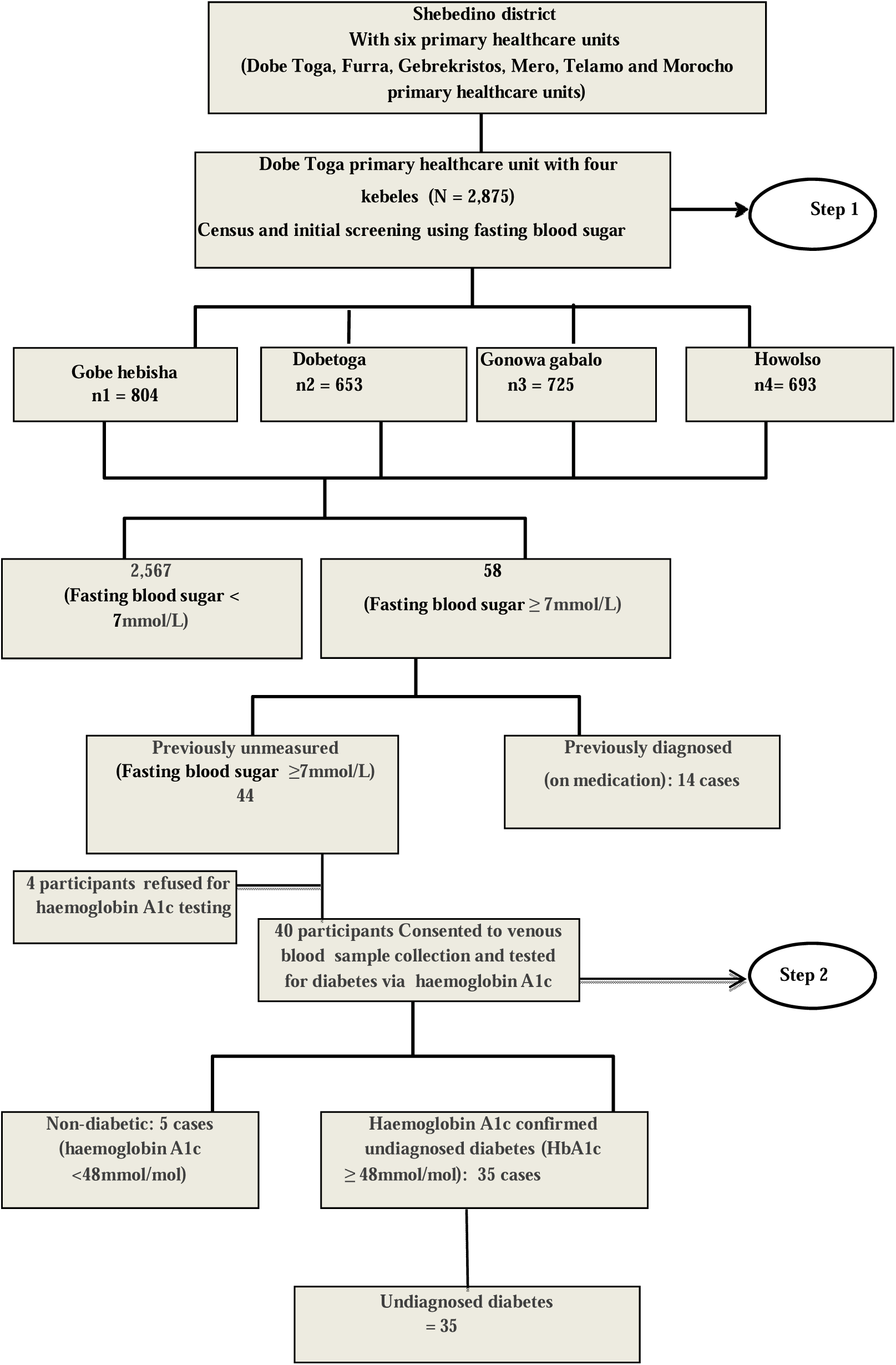
Study flow chart.

### Data collection tools and procedures

Data were collected using a pretested, structured, and interviewer-administered questionnaire based on the WHO STEPwise approach to surveillance for non-communicable diseases [32]. The questionnaire covered socio-demographic, economic, behavioural, and health-related factors, and was translated into *Sidaamu Affoo* and back-translated to ensure consistency.

Data collection was conducted by eight trained data collectors (two per kebele) with support from two laboratory technicians for blood sample collection. Two supervisors oversaw the field activities and quality assurance. Data collectors and supervisors were recruited based on their qualifications, experience, and language fluency, and received three days of training on study tools, Kobo Toolbox, and measurement techniques. The kobo Toolbox, an open-source platform, enabled mobile-based data collection. A pre-test on 5% of the sample was conducted in Remeda kebele, Hawella district, outside the main study site. Standardised procedures and equipment were used for physical and biochemical measurements; routine quality checks, including global position system (GPS) tracking and immediate data review, were carried out during data collection.

### Statistical Analysis

After data collection using Kobo Toolbox, the data sets were exported to Stata version 17 for Windows (StataCorp, College Station, USA) for analysis. Quantitative variables were cleaned and prepared through recoding, computation, and categorisation as necessary before analysis. For the outcome variable, blood samples were not obtained from four participants due to their unwillingness to provide them. These cases were excluded from the outcome analysis using listwise deletion. Given the very small proportion and the assumption of random missingness, no imputation was performed.

Descriptive analysis, including frequencies, percentages, means, and standard deviations, was used to summarize the characteristics of the study participants.

Bivariable linear regression analyses were performed to examine the unadjusted associations between each independent variable and the prevalence of diabetes identified via fasting blood sugar. Variables with p-values less than 0.25 in the bivariate linear regression were considered candidate variables for inclusion in the multivariable linear regression. Multivariable linear regression was then used to identify factors independently associated with fasting blood sugar levels. Multicollinearity among potential predictors was evaluated using the variance inflation factor (VIF), and linearity was assessed through a quantile-quantile (Q-Q) plot of the residuals. Results were presented as adjusted regression coefficients (β) with 95% confidence intervals (CI), and statistical significance was declared at p < 0.05.

## Result

### Description of study participants

A total of 2,875 study participants were involved in the study. Nearly half, 48.6% (1,394 of 2,875), of the study participants were aged between 45 and 54 years, with a median age of 55 years (IQR = 12 years). More than half, 55.1% (1,583 of 2,875), were females. Almost half, 51.6% (1,480 of 2,875), were housewives by occupation. Regarding literacy, 87.4% (2,513 of 2,875) could not read and write and 89.6% (1,772 of 2,875) had no formal education; see table 1. Among the 49 individuals identified with diabetes, 71.4% (35 of 49) were undiagnosed. Most diabetic cases were aged 45 to 54 years at 44.9% (22 of 49) and female at 63.3% (31 of 49); see table 2. One in ten, 10.2% (292 of 2,875), had ever smoked, and 19% (537 of 2,875) had ever consumed alcohol; see table 3.

**Table 1:** Socio-demographic and economic characteristics of study participants in Shebedino district, Sidama National Regional State, Ethiopia, 2025 (N=2,875).

| Variable | Category | Frequency | Percent |
| --- | --- | --- | --- |
| Age in years | 45–54 | 1,398 | 48.6 |
|  | 55–64 | 910 | 31.7 |
|  | 65+ | 567 | 19.7 |
| Sex | Male | 1,289 | 44.8 |
|  | Female | 1,586 | 55.2 |
|  | Others | 67 | 2.3 |
| Marital status | Currently not married | 322 | 11.2 |
|  | Currently married | 2,553 | 88.8 |
| Occupation status | Farmer | 1,353 | 47.1 |
|  | Housewife | 1,482 | 51.6 |
|  | Other* | 40 | 1.4 |
| Literacy | Can read and/or write | 362 | 12.6 |
|  | Cannot read and write | 2,513 | 87.4 |
| Education level | Attended primary or more | 300 | 10.4 |
|  | No formal education | 2,575 | 89.6 |
| Time to health facility | ≤ 30 minutes | 1,157 | 40.2 |
|  | > 30 minutes | 1,718 | 59.8 |
| Family size | ≤ 5 members | 2,115 | 73.6 |
|  | > 5 members | 760 | 26.4 |
| Wealth index | Poorest | 575 | 20.0 |
|  | Poorer | 600 | 20.9 |
|  | Middle | 550 | 19.1 |
|  | Richer | 648 | 22.5 |
|  | Richest | 502 | 17.5 |
| Estimated annual income | ≤ Quintal 1 | 1,031 | 35.9 |
|  | > Quintal, ≤ Quintal 2 | 1,147 | 39.9 |
|  | > Quintal 2, ≤ Quintal 3 | 482 | 16.8 |

Occupation: other\* includes daily laborer, merchant, governmental employees

**Table 2:** Distribution of undiagnosed and previously diagnosed diabetes among study participants in Shebedino district, Sidama National Regional State, Ethiopia, 2025

| Variable | Category | HbA1c<br>Undiagnosed<br>diabetes (n=35) |  | Previously<br>Diagnosed (n=14) |  | Total diabetic cases<br>(n=49) |  |
| --- | --- | --- | --- | --- | --- | --- | --- |
|  |  | n | % | n | % | n | % |
| Age in years | 45–54 | 18 | 51.4 | 4 | 28.6 | 22 | 44.9 |
|  | 55–64 | 7 | 20.0 | 4 | 28.6 | 11 | 22.5 |
|  | 65+ | 10 | 28.6 | 6 | 42.9 | 16 | 32.7 |
| Sex | Male | 14 | 40.0 | 4 | 28.6 | 18 | 36.7 |
|  | Female | 21 | 60.0 | 10 | 71.4 | 31 | 63.3 |
| Marital Status | Not married | 2 | 5.7 | 3 | 21.4 | 5 | 10.2 |
|  | Married | 33 | 94.3 | 11 | 78.6 | 44 | 89.8 |
| Occupation status | Farmer | 17 | 48.6 | 4 | 28.6 | 21 | 42.9 |
|  | Housewife | 18 | 51.4 | 10 | 71.4 | 28 | 57.1 |
|  | Other | 0 | 0.0 | 0 | 0.0 | 0 | 0.0 |
| Literacy | Can read/write | 4 | 11.4 | 0 | 0.0 | 4 | 8.2 |
|  | Cannot read/write | 31 | 88.6 | 14 | 100.0 | 45 | 91.8 |
| Education level | Primary or above | 2 | 5.7 | 0 | 0.0 | 2 | 4.1 |
|  | Not formally educated | 33 | 94.3 | 14 | 100.0 | 47 | 95.9 |
| Time to Facility | ≤ 30 minutes | 12 | 34.3 | 7 | 50.0 | 19 | 38.8 |
|  | > 30 minutes | 23 | 65.7 | 7 | 50.0 | 30 | 61.2 |
| Family Size | ≤ 5 members | 32 | 91.4 | 10 | 71.4 | 42 | 85.7 |
|  | > 5 members | 3 | 8.6 | 4 | 28.6 | 7 | 14.3 |
| Wealth Index | Poorest | 8 | 22.9 | 3 | 21.4 | 11 | 22.5 |
|  | Poorer | 5 | 14.3 | 4 | 28.6 | 9 | 18.4 |
|  | Middle | 5 | 14.3 | 2 | 14.3 | 7 | 14.3 |
|  | Richer | 8 | 22.9 | 3 | 21.4 | 11 | 22.5 |
|  | Richest | 9 | 25.7 | 2 | 14.3 | 11 | 22.5 |
| Annual Income | ≤ Quintal 1 | 10 | 28.6 | 1 | 7.1 | 11 | 22.5 |
|  | > Quintal, ≤ Quintal 2 | 14 | 40.0 | 7 | 50.0 | 21 | 42.9 |
|  | > Quintal 2, ≤ Quintile 3 | 7 | 20.0 | 4 | 28.6 | 11 | 22.5 |
|  | ≤ Quintal 1 | 4 | 11.4 | 2 | 14.3 | 6 | 12.2 |

**Table 3:** Behavioural and lifestyle characteristics of study participants in Shebedino district, Sidama National Regional State, Ethiopia, 2025 (N = 2,875).

| Variable | Category | Frequency | Percent |
| --- | --- | --- | --- |
| Smoking history cigarette | No | 2,583 | 89.8 |
|  | Yes | 292 | 10.2 |
| Family member smokes tobacco | Yes | 50 | 1.7 |
|  | No | 2,825 | 98.3 |
| Friends smoke tobacco | Yes | 93 | 3.2 |
|  | No | 2,782 | 96.8 |
| Alcohol drinking history | No | 2,338 | 81.3 |
|  | Yes | 537 | 18.7 |
| Khat chewing history | No | 2,126 | 74 |
|  | Yes | 749 | 26 |
| Eat fruit between meals | Yes | 1,333 | 46.4 |
|  | No | 1,542 | 53.6 |
| Eat vegetables between meals | Yes | 1,843 | 64.1 |
|  | No | 1,032 | 35.9 |
| Drink coffee | Yes | 2,793 | 97.2 |
|  | No | 82 | 2.8 |
| Vigorous-intensity activity | Yes | 1,340 | 46.6 |
|  | No | 1,535 | 53.4 |
| Moderate-intensity activity | Yes | 749 | 26.1 |
|  | No | 2,126 | 73.9 |

The mean waist-to-height ratio among the study participants was 0.44 (95% CI: 0.43-0.44), and the mean waist circumference was 68.9 cm (95% CI: 68.6, 69.2 cm). The mean body weight was 47.0 kg (95% CI: 46.7, 47.3 kg), and the mean body mass index of the study participants was 18.9 kg/m^2^ (95% CI: 18.8, 19.0 kg/m^2^). Regarding nutritional status, nearly 46.2% (1,327 of 2,875) were underweight, 50.9% (1,463 of 2,875) had a healthy weight, and only 3% (85 of 2,875), were categorised as overweight.

### Prevalence of undiagnosed diabetes

The prevalence of undiagnosed diabetes mellitus confirmed using HbA1c was 1.2% (35 of 2,871; 95% CI: 0.9% - 1.7%) of the study participants. Previously diagnosed diabetes was identified in 0.5% (14 of 2875; 95% CI: 0.3% - 0.8%) of the study participants. Among the 14 diabetic patients, 71.4% (10 of 14) were under treatment with metformin and 28.6% 4 of 14) were under treatment with insulin. Overall, the total diabetes prevalence confirmed by HbA1c results or prior diagnosis (based on fasting blood sugar and verification of diabetes medications or packaging) was 1.7% (49 of 2,871; 95% CI: 1.3% -2.3%). The prevalence of elevated fasting blood sugar based on fasting blood sugar alone was 2.1% (59 of 2,875; 95% CI: 1.6% - 2.6%).

### Predictors of fasting blood sugar (mmol/L)

After adjusting for other variables in the multivariable linear regression model, age, estimated annual income in kind, and waist-to-body mass index (Waist-to-BMI) were significantly associated with elevated fasting blood sugar measured in mmol/L. Individuals aged 65 and above had a 0.18 mmol/L higher fasting blood sugar level compared to those aged 45-54 (β = 0.18; 95% CI: 0.06, 0.30, p = 0.004). Study participants with estimated annual income earning of two to three quintiles had a 0.14 mmol/L higher fasting blood sugar compared to the lowest income category (≤ Quintile 1) (β = 0.14; 95% CI: 0.01, 0.16; p = 0.032); see table 4.

**Table 4:**
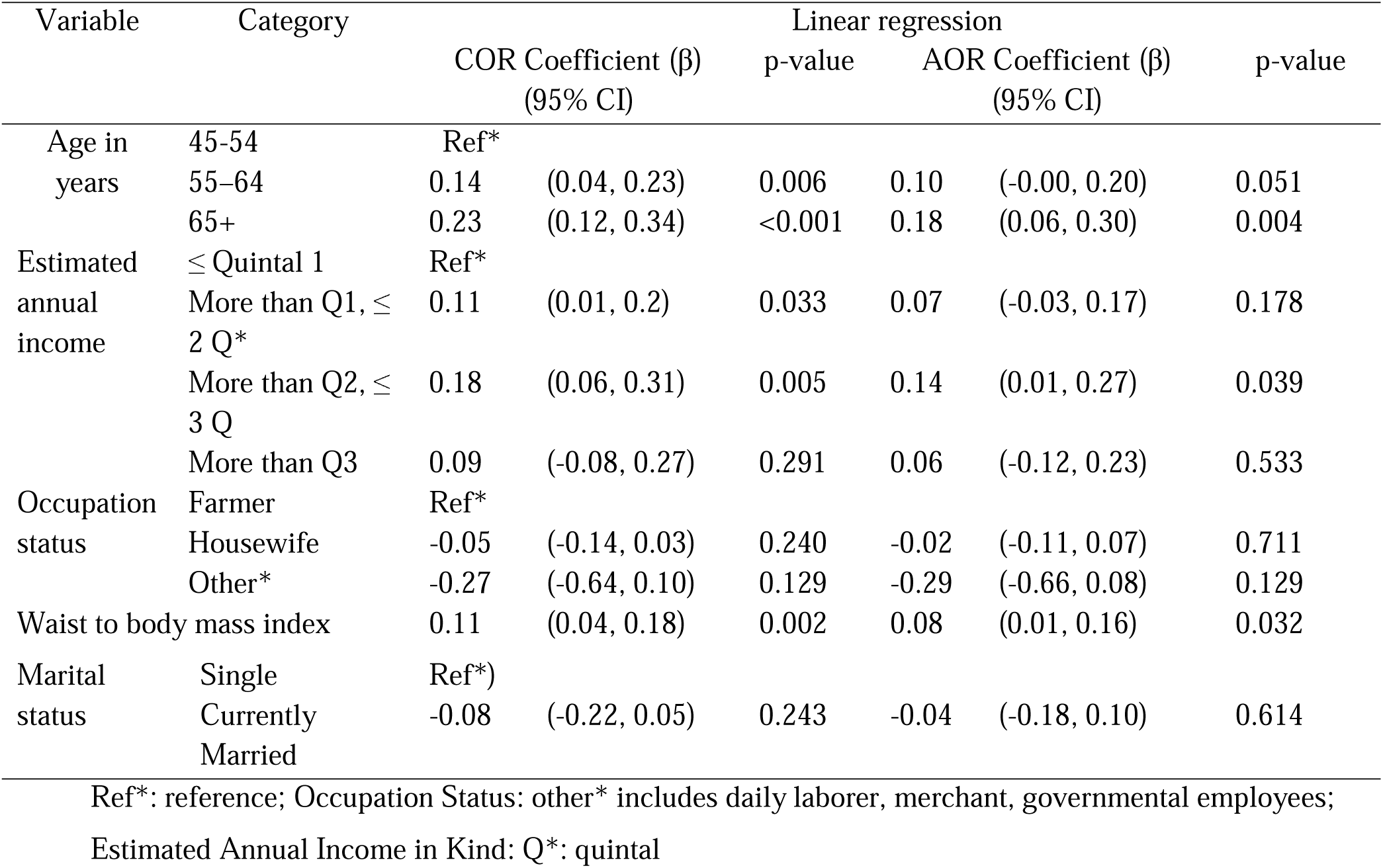
Predictors of fasting blood sugar among study participants in Shebedino district, Sidama National Regional State, Ethiopia, 2025 (N=2,875).

## Discussion

This community-based cross-sectional study, conducted in rural Sidama, Ethiopia, employed a two-step diagnostic approach combining fasting blood sugar screening with confirmatory haemoglobin A1c testing. The prevalence of undiagnosed, previously diagnosed, and total diabetes was found to be low. In this study, age, estimated annual income, and waist-to-body mass index were significantly associated with fasting blood sugar measured in mmol/L.

The prevalence of undiagnosed diabetes (1.2%) observed in this study is lower than global estimates, where nearly one in ten adults (10.5%) is affected, and also the African regional estimate of 4.5% [4]. Significantly higher burdens have been reported in Asia, such as Pakistan: 23.9%, and India: 19.2% [33–35], and urban Africa, such as Mali: 7.5% [36], Uganda: 7.4% [37]. This difference might be due to the methodological differences; many high-prevalence studies rely on fasting blood sugar alone, and contextual factors such as protective rural lifestyles, higher physical activity, and unprocessed diets in this study setting [38].

Compared to the national and regional estimates from previous Ethiopian studies, the observed prevalence remains lower. For example, WHO (2015) estimated a national prevalence of 3.2% [39], while a 2019 meta-analysis reported a pooled prevalence of 6.5% [13]. Urban studies have consistently shown higher prevalence, such as in western Ethiopian towns (5.6%) [40], Dessie in northern Ethiopia (6.8%) [41], Hossana town in southern Ethiopia (5.7%) [42], Mizan town in southern Ethiopia (8.1%) [43], Hawassa (14.4%) [14], northern Ethiopia (6.5%) [44], all of which primarily used fasting blood sugar alone for diagnosis.

The lower prevalence observed in this study may be due to both the diagnostic method and the rural settings. Unlike previous studies that relied solely on fasting blood sugar, which can capture transient hyperglycaemia and potentially overestimate prevalence, this study applied a two-step approach: fasting blood sugar screening followed by confirmatory haemoglobin A1c testing, a more stable marker of chronic glycaemia. Moreover, earlier studies were conducted in urban areas where sedentary lifestyles and processed food consumption are more common. In contrast, the present study population was predominantly rural, characterised by limited urbanisation, reduced access to processed foods, and higher physical activity, all of which may contribute to a lower diabetes risk [13, 45].

Undernutrion may provide a possible explanation for the observed low prevalence of undiagnosed diabetes mellitus in the study population. Nearly half of the study participants were classified as underweight, suggesting a widespread lean body composition attributed to chronic caloric deficiency and high levels of physical activity levels. This metabolic profile characterised by reduced fat stores and lower insulin resistance has been associated with a decreased risk of developing diabetes. Individuals experiencing long–term malnutrition often exhibit lower body mass index, albumin, haemoglobin, triglyceride, and high-density lipoprotein, while paradoxically showing elevated glycosylated haemoglobin levels [6–8]. Moreover, haemoglobin A1c may underestimate true diabetes prevalence in lean populations or those affected by haemoglobinopathies [1]. Survivor bias could further play a role, as individuals with undiagnosed or poorly managed diabetes may experience in premature mortality in resource-limited settings, thereby reducing the apparent prevalence of the disease [46].

This finding aligns with other Ethiopian studies, such as those from Gondar and Jimma, which reported that people with diabetes often have lower body mass index and are typically subsistence farmers or unemployed, suggesting a link between diabetes and undernutrition in rural settings [47]. Supporting evidence has documented signs of malnutrition in individuals with diabetes, such as reduced mid-upper arm circumference and low body mass index [48]. Notably, such individuals often present features that differ from classic type 2 diabetes, including body mass index below 18.5 kg/m^2^, resistance to ketosis, and low serum C-peptide levels. These unique clinical characteristics have led to debate over whether undernutrition-related diabetes represents a distinct disease entity, potentially warranting separate recognition in the WHO classification [49].

In this study, individuals aged 65 and above had significantly higher fasting blood sugar compared to those aged 45-54, consistent with previous findings showing that advanced age is a key risk factor for impaired glucose metabolism [13,50–52]. As people age, glucose tolerance, insulin sensitivity, and insulin secretion typically decline [53]. Estimated annual income was also significantly associated with fasting blood sugar levels, in line with findings from rural Bangladesh [54]. Rising income in rural communities may contribute to the early stages of nutrition transition, leading to higher consumption of energy-dense processed foods and reduced physical activity [55, 56]. In contrast, individuals with very low income often engage in physically demanding daily labour, which may offer some protection against hyperglycaemia [57].

Waist-to-body mass index was also significantly associated with fasting blood sugar levels. This supports evidence that central obesity, measured by waist-to-body mass index or waist-to-hip ratio, is strongly linked to higher fasting blood sugar through its effect on insulin resistance, chronic inflammation, and metabolic disorder [58–60].

The study has several strengths. It is among the few large, community-based cross-sectional studies in rural Ethiopia targeting adults aged 45 and above, offering a valuable epidemiological picture of diabetes. The two-step diagnostic approach using both fasting blood sugar and HbA1c testing improves accuracy; unlike most prior Ethiopian studies rely only on fasting blood sugar, which may overestimate prevalence. HbA1c provides a more stable measure of chronic hyperglycaemia.

However, findings should be interpreted with caution. The predominantly rural, homogenous sample may limit generalizability. Self-reported lifestyle behaviours (tobacco use, alcohol consumption, and khat chewing) may be subject to recall bias and social desirability bias, though the WHO-Stepwise tools and trained data collectors helped to mitigate this. Unmeasured conditions such as anaemia may affect the accuracy of HbA1c reading, but the two-step diagnostic approach aimed to reduce the potential misclassification. The absence of data on diet, physical activity, and genetics may confound results, though detailed socio-demographic and behavioural variables were adjusted for. The cross-sectional design also limits causal inference.

These results have important implications for rural health planners, clinicians, and policymakers, who should prioritise community-based screening, early detection, and interventions that maintain healthy diets and physical activity.

## Conclusion

The study found a low prevalence of undiagnosed diabetes using a two-step diagnostic method. Elevated fasting blood sugar was associated with age, estimated annual income, and waist-to-body mass index.

To maintain this low prevalence, community screening and health education should target older adults, households with increasing incomes, and individuals with higher waist-to-body mass index. Local health services should promote a healthy diet, physical activity, and regular blood sugar monitoring.

Further research should explore whether undernutrition lowers diabetes risk or if protective lifestyles such as physical activity, limited processed food intake, and lean body composition are protective factors for lower fasting blood sugar. Studies should distinguish forms of undernutrition and investigate metabolic markers like insulin sensitivity and beta-cell function, particularly in lean individuals with atypical diabetes. Research should also assess whether undernutrition-related diabetes constitutes a distinct clinical subtype. Longitudinal and intervention studies are needed to understand how evolving nutrition, income, and lifestyle patterns in rural communities affect diabetes risk. Urban-rural comparisons using standardised diagnostics could improve national estimates and guide context-specific public health strategies.

### Data availability statement

The data supporting this study’s findings are openly available as supplementary material, including the anonymised data set (S1), structured questionnaire (S2), study flow diagram (S3), and the STROBE checklist (S4). All supplementary files can be accessed with this manuscript submission.

## Data Availability

All data produced in the present study are available upon reasonable request to the authors

## Ethics Statements

### Consent for publication

Not applicable

### Ethics approval

The study was reviewed and approved by the Institutional Review Board (IRB) of Hawassa University, College of Medicine and Health Sciences, which issued an ethical clearance letter (Ref No. IRB/100/16, dated 12/03/2024. An official support letter was taken from the Sidama National Regional State Public Health Institute. Permission was also granted from the Shebedino district health office and the kebeles’ administrators.

During sample collection and interviewing, study participants were informed about the purpose of the study, and written consent was obtained. For individuals unable to read or write, the consent was read aloud, and a thumbprint was collected to document agreement. Confidentiality was maintained, with participant names kept anonymous, and the tool password-protected. Counselling and health education were provided for those diagnosed with diabetes, and they were linked to Dobe Toga Primary Healthcare unit for follow-up.

## Acknowledgments

We are indebted to the Government of Norway for funding this research through the NORHED SENUPH-II program. We also extend our acknowledgement to Shebedino district for facilitating the research, as well as the data collectors, and the study participants for their full engagement.

## Author Contributions

Desalegn Tsegaw Hibstu, Melaku Haile Likka, Hiwot Abera Areru, Betelhem Eshetu, and Bernt Lindtjørn contributed to the conceptualisation, data curation, formal analysis, investigation, methodology, project administration, resources, software, supervision, validation, and visualization of the study. All authors wrote the original draft and contributed to the review and editing of the manuscript. All authors read and approved the final version.

## Patient and public involvement

Patients or the public were not involved in the design, or conduct, or reporting, or dissemination plans of our research.

